# Napsin A-specific T cell clonotypes are associated with improved clinical outcomes in patients receiving checkpoint immunotherapy for metastatic non-small cell lung cancer

**DOI:** 10.1101/2025.03.10.25323586

**Authors:** Natalie J. Miller, Christina S. Baik, Joel W. Neal, Fangdi Sun, Rafael Santana-Davila, Sylvia Lee, Keith D. Eaton, Renato G. Martins, Cristina Rodriguez, Heather A. Wakelee, Sukhmani K. Padda, Eric Q. Konnick, Alex Camai, Tatyana Pisarenko, Viswam S. Nair, A. McGarry Houghton, Shin-Heng Chiou, Diane Tseng

## Abstract

**Background:** Napsin A is normally expressed in human lung pneumocytes and is a highly expressed cancer antigen in lung adenocarcinoma. We examined whether T cells specific for Napsin A may play a role in immune checkpoint inhibitor (ICI)-mediated responses. We utilized bulk TCR repertoire data to assess whether the presence of Napsin A-specific clonotypes in the peripheral blood was associated with improved clinical responses to ICI.

**Methods:** Patients with metastatic non-small cell lung cancer (NSCLC) receiving anti-PD-(L)1 (alone or in combination) were enrolled at Fred Hutchinson Cancer Center and Stanford University Medical Center (n=62; histology of adenocarcinoma n=48, squamous n=9, NSCLC/other n=5). Peripheral blood mononuclear cells (PBMC) were collected for genomic DNA isolation pre- and post-treatment (range 3 weeks - 3 months). TCRβ was bulk sequenced via the immunoSEQ platform (Adaptive Biotechnologies). Napsin A-specific TCRβ sequences were identified from publicly available data and their frequencies were quantified in each patient sample. We examined whether overall survival (OS) and progression-free survival (PFS) outcomes differed in patients with or without detectable Napsin A-specific TCRs (herein Napsin TCRs). We used Cox proportional hazards regression to assess the association between detectable Napsin TCRs and PFS or OS in univariable and multivariable analyses.

**Results:** Napsin TCRs were detectable in the blood in a large fraction of our cohort (n=25/62 [40%] [pre-treatment; n=21/42 [50%] post-treatment). Patients with detectable Napsin TCRs had a significant improvement in OS compared to patients without these TCRs (median OS 45.4 vs 14.8 months, p=0.0043 pre-treatment; median OS 55.4 vs 18.9 months, p=0.0066 post-treatment). Among 27 *HLA-A*02* carriers of 55 HLA-typed patients (49%), patients with detectable pre-treatment Napsin TCRs had a significant improvement in OS (median 60.2 vs 16.5 months, p=0.0054) and PFS (median 21.5 vs. 7.2 months, p=0.031) compared to patients without these TCRs. In univariate and multivariate analysis, the presence of Napsin TCRs pre-treatment was associated with improved OS (p=0.0057, HR 0.40, 95% CI 0.21-0.76 univariate; p=0.033 HR 0.45, 95% CI 0.23-0.91 multivariate).

**Conclusions:** Napsin TCRs are frequently detected in patients with NSCLC and are associated with improved OS in patients with NSCLC receiving ICI.

**KEY MESSAGES:** *What is already known on this topic:* Whether T cell immune responses against non-mutated tumor antigens play a role in checkpoint immunotherapy responses remains largely unknown.

*What this study adds:* Using a multicenter cohort of patients with advanced NSCLC on ICI we demonstrate that presence of TCRs specific for the lung adenocarcinoma tumor antigen Napsin A at pre- or early post-treatment timepoints is associated with improved overall survival (OS). This work is novel in showing that an overexpressed non-mutated proteins elicits specific T cells that are correlated with response to ICI.

*How this study might affect research, practice or policy:* T cells recognizing the self-antigen Napsin A may play a role in checkpoint immunotherapy responses. This suggests that T cells recognizing overexpressed non-mutated antigens may shape clinical outcomes to checkpoint immunotherapy.

## INTRODUCTION

The antigen specificities of T cells that are key to the success and durability of checkpoint immunotherapy remain largely unknown. To date, much work has focused on neoantigen-specific T cells ^1–4^, but whether T cells targeting self-antigens like Napsin A play a role in ICI response remains unknown. T cell receptor (TCR) sequencing is now widely used in translational research to comprehensively evaluate patients’ immune repertoires, but there have been few studies focusing on antigen-specific T cell responses, and most studies have focused on clonal dynamics in an antigen-agnostic approach ^5,6^. These limitations are largely due to the challenges of interrogating genetically diverse cancers and immunologically diverse human leukocyte antigen (HLA) contexts.

Napsin A is a tumor-associated self-antigen highly expressed in NSCLC ^7^. It is an aspartic proteinase normally expressed in Type 2 pneumocytes of the lung ^8^. Detection of Napsin A by immunohistochemistry (IHC) is strongly associated with adenocarcinoma and is routinely used as a clinical diagnostic to differentiate adenocarcinoma from squamous cell carcinoma of the lung ^9^, as well as to differentiate adenocarcinoma originating from lung versus another primary organ. In a recent study of checkpoint pneumonitis, Napsin A-specific CD8+ T cells emerged as an autoreactive T cell population that was enriched in tumors and inflamed lung relative to the blood ^10^. Napsin A-specific CD8+ T cells from their study were isolated from *HLA-A*02+* patients and their T cell receptor beta chain (TCRβ) sequences made publicly available.

If Napsin A-specific T cells could mediate anti-tumor effects in patients with NSCLC treated with immune checkpoint inhibitors (ICI), we hypothesized that the presence of Napsin A-specific clonotypes would be associated with improved clinical benefit from ICI. In this study we sequenced the bulk TCRβ repertoire from PBMC of 62 patients with metastatic NSCLC treated with anti-PD-1 at pre- and post-treatment timepoints. We determined the incidence and frequency of the 20 previously reported Napsin A-specific TCRβ clonotypes ^10^ in each sample and whether presence of Napsin TCRs were associated with improved outcomes to ICI in treated patients.

## METHODS

### Patients

Patients were enrolled from either Fred Hutchinson Cancer Center (FHCC) or Stanford University Medical Center after approval by respective institutional review board and ethics committee. Written informed consent was obtained from all patients. All research was performed in accordance with relevant guidelines and regulations, including the Declaration of Helsinki. Patients were enrolled from Sept 2013 to July 2020 at FHCC and Nov 2017 to Feb 2019 at Stanford. Study eligibility for patients included diagnosis of metastatic NSCLC and treatment with anti-PD-(L)1, alone or in combination with additional immunotherapy, with chemotherapy, and/or with investigational agents (specifics in Results section). Patients were excluded for lack of clinical response data or molecular alteration in genes such as *EGFR*, *ALK* or *ROS1*, where mechanisms of resistance to immunotherapy are distinct. Peripheral blood mononuclear cells (PBMCs) were collected at pre- and post-treatment (range 3 weeks - 3 months) and viably cryopreserved. Clinical data was obtained by retrospective chart review through data cut off of March 15^th^, 2024. Important pre-treatment clinical and demographic data included gender, race, histology, smoking history, Eastern Cooperative Oncology Group (ECOG) performance status, prior therapies, and laboratory studies at time of treatment start. Durable clinical benefit (DCB) was defined as ongoing clinical benefit at 6 months after initiating ICI by the treating physician. Development of immune related adverse events were identified by the treating physician.

### T cell receptor **β** profiling

Genomic DNA was extracted from frozen PBMC via DNA Blood kit (Qiagen) and submitted for T cell receptor β chain (*TRB)* variable region sequencing via the immunoSEQ hsTCRB v3.0 assay based on recommended sample guidelines ^11^ (Adaptive Biotechnologies, Seattle, WA). We adopted all TCR metrics that were derived from the immunoSEQ portal by default and included only productive TCR templates for all downstream analyses. TCR repertoire metrics including TCR richness (defined as number of unique productive rearrangements ^11^), productive clonality, fraction of T cells of all nucleated cells, Shannon diversity index, and frequency of clones present at >0.01% were analyzed applying relevant features from immunoSEQ® 3.0 Analyzer (Adaptive Biotechnologies, Seattle WA USA). Clonality was defined as 1-Peilou’s evenness ^12^. Shannon index (entropy) was employed to measure the diversity of TCR clonotypes within a sample ^13^. Morisita-Horn index was performed to compare similarity between samples as previously described ^14^. Comparisons of these metrics among the cohort was performed using custom R code (R Foundation for Statistical Computing, Vienna, Austria). The TCR sequencing data is deposited in the Adaptive Biotechnologies Published Projects Database and can be accessed at: https://clients.adaptivebiotech.com/pub/miller-2025-s.

Publicly available TCRβ Complementarity-determining region 3 (CDR3) sequences specific for Napsin A (n = 20) were obtained ^10^; similarly, TCRβ CDR3 sequences specific for epitopes from influenza, Epstein–Barr virus (EBV), Cytomegalovirus (CMV), melanoma-associated antigen recognizing T cells (MART-1) and New York esophageal squamous cell carcinoma 1 (NY-ESO-1) were obtained from public databases ^15–17^ (complete list of >8000 TCRβ upon request). The summed frequency of the TCRβ sequences specific for each antigen was calculated for each sample. In subsequent analyses, patients were stratified by having TCR frequency above or below the median. For Napsin A- and NY-ESO-1, the median value was 0 and therefore above median equaled “detectable”. In addition, the mean frequency and SD of each antigen-specific TCR was calculated among patients with or without DCB (**Table 2**); one-sided Wilcoxon rank sum tests were used to assess difference of the means given the assumption of higher TCR frequency for patients with DCB.

### HLA genotyping

Genomic DNA was extracted from patient PBMC followed by DNA quantification (Qiagen). HLA genotyping was completed via either ScisGo Genetics, Geraghty Lab (Fred Hutchinson Cancer Center, Seattle, WA), or Histogenetics (Ossining, NY).

### Tumor mutational profiling

Clinical standard-of-care tumor mutational data was available for 54 of 62 patients; typically from the initial diagnostic sample. Most tumors from patients within the FHCC cohort were assayed using the UW-OncoPlex^TM^ assay, a multiplexed mutation assay for tumor tissue that assesses all classes of mutations in >400 genes related to cancer treatment, prognosis, or diagnosis ^18^; other patients had clinical testing per treating physician standard of care (i.e., Stanford Actionable Mutation Panel for Solid Tumors, Foundation One). Clinically relevant mutations including those in *EGFR*, *ALK*, *ROS1*, *KRAS*, *MET*, *RET*, *BRAF*, *ERBB2*, and/or *NTRK1/2/3* were reported.

### Statistical analysis

Descriptive statistics of the clinical and demographic data included median with interquartile range between 25^th^ and 75^th^ percentiles for numerical variables as well as percentages for categorical variables. Tests for independence were performed using Chi squared and two-sided Fisher’s exact tests depending on sample size. Patients were stratified as having detectable versus undetectable Napsin TCRs for PFS and OS analysis. PFS was defined as time from ICI start to radiographic disease progression, defined by treating physician. OS was defined as time from ICI start to death. Patients who did not experience the endpoint of progression or death were censored at date of last follow up. Cox proportional hazards regression was used to assess the association between PFS or OS and clinical characteristics plus TCR metrics including detectable Napsin TCRs in univariable and multivariable analyses. Kaplan–Meier method was used to estimate the PFS and OS functions, and log-rank test was used to assess difference. Comparisons of patients with DCB vs non-DCB (NDCB) were performed using one-sided Wilcoxon test with the assumption that DCB associates with greater values. Kolmogorov-Smirnov and Shapiro-Wilk tests were used to exclude the possibility of normality in data distribution.

## RESULTS

### Patient demographics

Patients with metastatic NSCLC receiving anti-PD-(L)1 (alone or in combination) were enrolled at Fred Hutchinson Cancer Center (n = 53) and Stanford University Medical Center (n = 19). In this cohort, patients were excluded for lack of clinical response data (n = 4) or molecular alteration in genes such as *EGFR*, *ALK* or *ROS1*, where mechanisms of resistance to immunotherapy are distinct (n = 6) resulting in 46 evaluable patients from FHCC and 16 from Stanford. Pre-treatment PBMC were available for the entire cohort (n=62) while matched pre- and post-treatment PBMC were available for 42 patients (**Figure 1**). Baseline clinical data including age, gender, smoking history, tumor histology, PD-L1 IHC expression, ECOG performance status, and line of therapy was collected and summarized in **Table 1**. Histology for the majority of the cohort was adenocarcinoma (n = 48), followed by squamous (n = 9), and poorly differentiated NSCLC (n = 5). ICI was given as 1^st^ line in 63% of patients (n = 39), 2^nd^ line (n = 13), 3^rd^ line (n = 7) or 4^th^ line and beyond (n = 3). Type of anti-PD-(L)1 included pembrolizumab in 74% (n = 46), atezolizumab (n= 8, 13%), nivolumab (n= 6, 9.7%), durvalumab (n= 1) and avelumab (n=1). Anti-PD-(L)1 was given alone (n= 34), with chemotherapy (n=23, predominantly carboplatin plus pemetrexed), with investigational agents (n=3; plinabulin n=2, mocetinostat n=1), or as combination immunotherapy (n=2; with anti-4-1BB in n=1 and anti-CTLA-4 in n=1).

**Figure 1:**
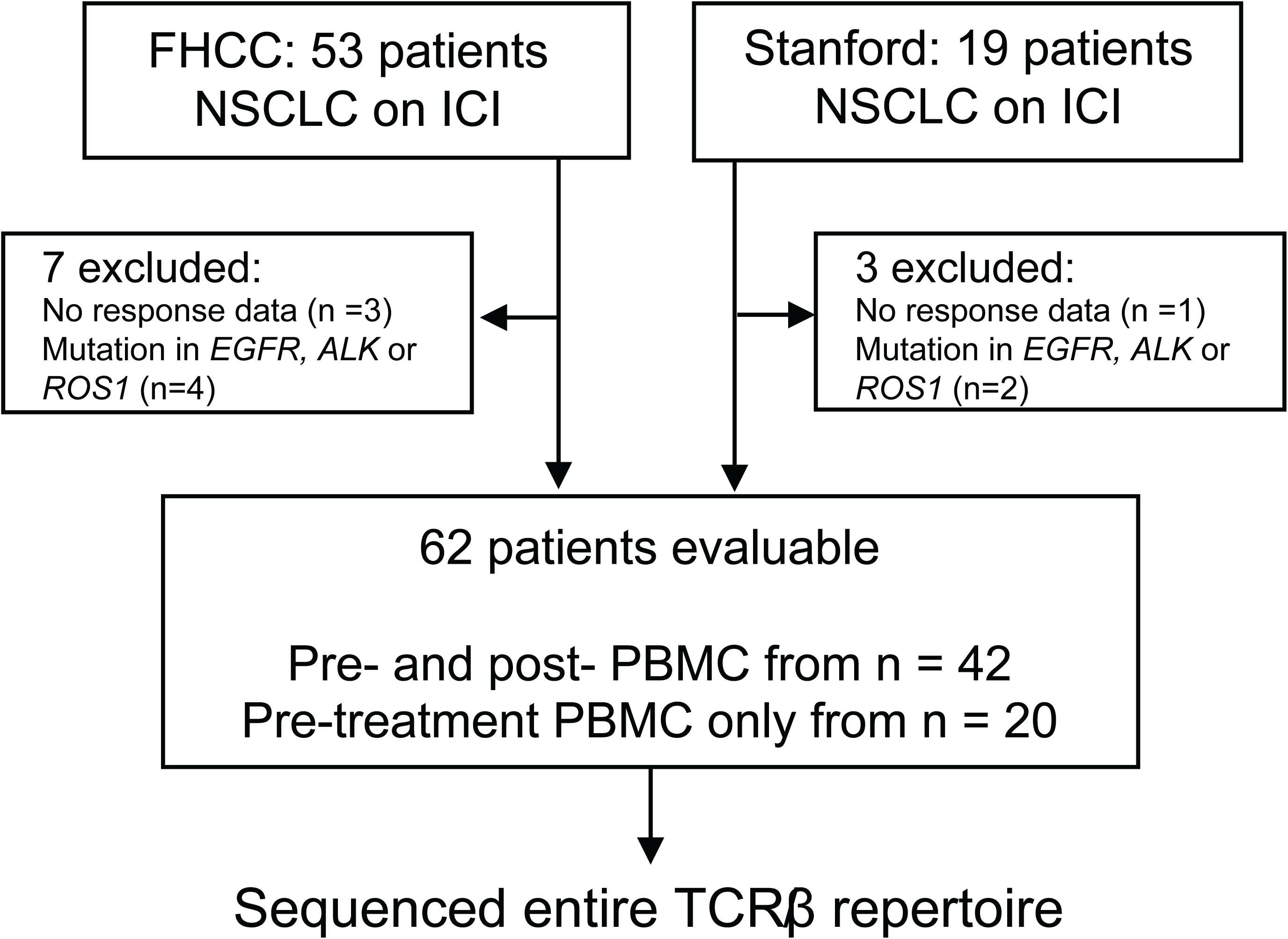
Consort flow diagram for study participants. Targetable mutation included EGFR E19del (n = 4) or ALK fusion (n = 2). Post-treatment PBMC obtained between 1 - 3 months after start of ICI for FHCC cohort and at 3 weeks for Stanford cohort.

**Table 1:** Baseline Clinical Characteristics.

| **Table 1: Baseline Clinical Characteristics** |  |
| --- | --- |
| **Characteristic** | **n = 62** |
| **Age (years), Median (Range)** | 71 (38 - 89) |
| **Male sex, n (%)** | 26 (42%) |
| **Former/current smoker, n (%)** | 54 (87%) |
| **Histology, n (%)** |  |
| adenocarcinoma | 48 (77%) |
| squamous | 9 (15%) |
| poorly differentiated | 5 (8%) |
| **PD-L1 %, n (%)** |  |
| < 50% | 31 (50%) |
| ≥ 50% | 20 (32%) |
| unknown | 11 (18%) |
| **ECOG, n (%)** |  |
| 0 | 12 (19%) |
| ≥ 1 | 50 (81%) |
| **Line of Therapy, n (%)** |  |
| 1<sup>st</sup> | 39 (63%) |
| 2<sup>nd</sup> and beyond | 23 (37%) |
| **HLA A\*02 carrier, n (%)** |  |
| Positive | 27 (44%) |
| Negative | 28 (45%) |
| Unknown | 7 (11%) |

### Patient outcomes to ICI

The median follow-up time for all patients in this cohort was 6.9 years (95% CI 5.9 – 7.3 years) by reverse Kaplan Meier method. Half of patients (n=31) experienced durable clinical benefit (DCB), which was defined as ongoing clinical benefit at 6 months.

### TCR repertoire characteristics among patients with and without DCB

To gain insights into the relationship between descriptive metrics of the T cell repertoire and clinical response to treatment, we analyzed TCR richness, productive clonality, Shannon’s diversity index, the frequency of TCR clones above 0.01% of the repertoire and compared these values among patients with or without DCB to ICI (**Supplemental Figure 1**). In addition, we measured the Morisita-Horn index of repertoire similarities between pre- and post-treatment samples (n=42) among patients with and without DCB to ICI. There was significantly higher TCR richness among patients with DCB compared to NDCB (one-sided p=0.001) at pre-treatment timepoint (n=62), which was not significant post-treatment (n=42, p=0.051, Wilcoxon rank sum test). Shannon’s diversity index was also higher among patients with DCB compared to NDCB at pre-treatment (p=0.012) which was not significant in post-treatment samples (p=0.065). There was no significant difference in clonality or percentage clones above 0.01% among patients with DCB compared to NDCB at either pre-or post-treatment timepoints, and no significant difference in Morisita-Horn index among patients with or without DCB (p=0.71). These findings generally align with published reports of TCR repertoire analyses in lung cancer ^19–21^, acknowledging some variation across studies.

### Napsin TCRs are detectable in the peripheral blood of NSCLC patients

To accommodate the idiosyncratic nature in TCR clonal frequencies, we defined patients with evidence of T cell responses against Napsin A as having any detectable Napsin TCRs in the blood. We identified one or more of the 20 publicly available Napsin TCRs in the blood in a large fraction of our cohort (n=25 out of 62 at pre-treatment; n= 21 out of 42 at post-treatment timepoint). Napsin TCRs were found both among patients from FHCC (n= 20 of 46 pre-treatment, n=14 of 26 post-treatment) and Stanford (n= 5 of 16 pre-treatment, n= 7 of 16 post-treatment).

### Improved OS among patients with detectable Napsin TCRs in the peripheral blood

We next performed survival analysis among patients with or without detectable Napsin TCRs at pre- or post-treatment time points (**Figure 2**). There was no significant difference in PFS between patients with detectable or not detectable Napsin TCRs in pre-treatment (13.1 months for Napsin TCR detectable vs 5.2 months for Napsin TCR undetectable; p= 0.16) or post-treatment (15.0 months for Napsin TCR detectable vs 6.2 months for Napsin TCR undetectable; p=0.34) PBMCs. However, having detectable Napsin TCRs was associated with improved OS. Specifically, patients with identifiable Napsin TCRs within PBMC from the pre-treatment timepoint (n=25) had significantly better OS (median 45.4 months) compared to those without (median 14.8 months, n=37, p=0.0043, HR 0.40). In post-treatment PBMC, there was similarly better OS among patients with detectable Napsin TCRs (median 55.4 months) compared to patients without detectable Napsin TCRs (median 18.9 months, p= 0.0066, HR 0.35). Given that Napsin A protein expression by IHC is associated with adenocarcinoma histology, we assessed whether the frequency of Napsin TCRs varied by NSCLC histology. The mean frequency of Napsin TCR was higher in patients with adenocarcinoma (1.9 × 10^−3^ %, n=48) compared to squamous cell carcinoma (SCC) (5.9 × 10^−4^%, n=9), although not statistically significant by Wilcoxon rank sum testing. Next, we performed exploratory analysis to compare OS among patients with adenocarcinoma versus SCC, stratified by presence of Napsin TCRs (**Supplemental Figure 2**). Among patients with adenocarcinoma, patients with detectable Napsin TCRs (n=19) had improved OS compared to patients without Napsin TCRs (n=29, p= 0.0019, HR 0.30). In contrast, SCC patients with detectable Napsin TCRs (n=4) did not have significantly different OS compared to those without Napsin TCRs (n=5, p=0.90). While the number of SCC cases was limited, OS in these patients was similar to adenocarcinoma patients without detectable Napsin TCRs.

**Figure 2:**
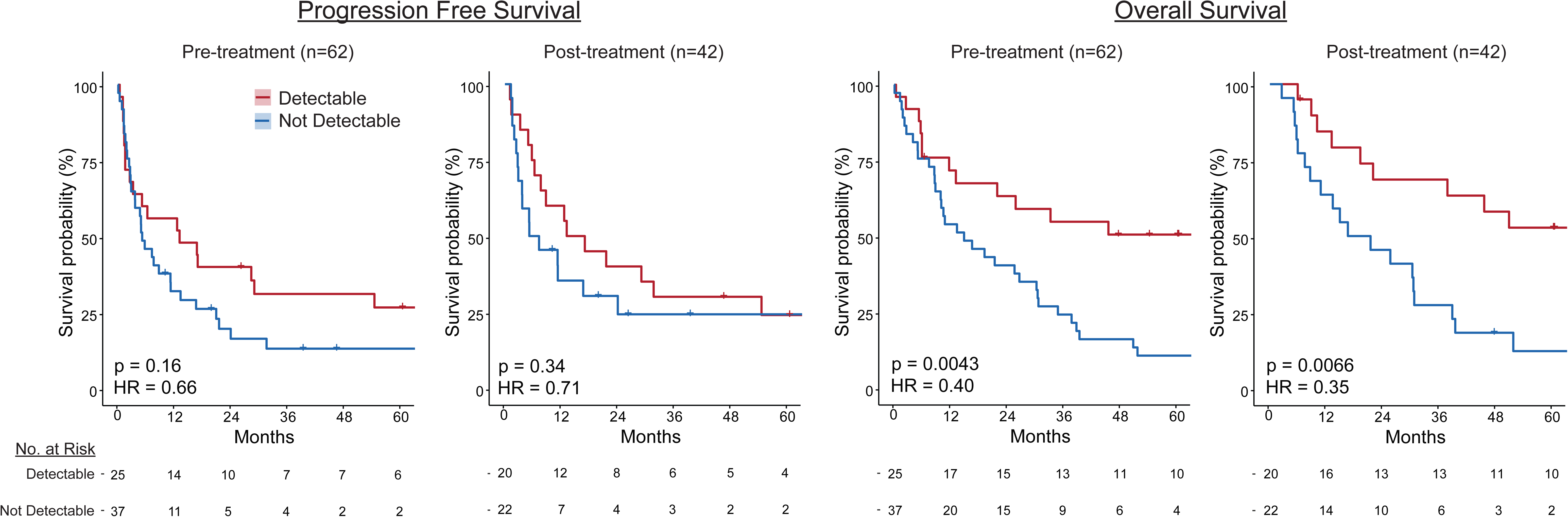
Patients with detectable Napsin TCRs have improved OS, but not PFS, when measured from both pre- and post-treatment PBMC. Kaplan-Meier method used to plot survival curves for patients with detectable Napsin TCRs (red) or without detectable Napsin TCRs (blue) in PBMC from pre- or post-treatment timepoints. Both PFS and OS calculated from time of treatment start. Number at risk shown below each plot. Log-rank test used for comparisons.

### Improved PFS and OS among patients with *HLA-A*02* and detectable Napsin TCRs

The 20 Napsin TCRs analyzed in our study were derived from four patients carrying at least one *HLA-A*02* allele (*A*02* carriers) ^10^, suggesting that many of them are likely A*02 restricted. Next, we assessed whether these Napsin TCRs were enriched and/or further associated with improved survival among *A*02* carriers. We performed genotyping for HLA class I alleles on 55 patients in our cohort with available remaining genomic DNA after TCR sequencing (**Supplemental Table 1**) and identified 27 of the 55 patients (49%) as *A*02* carriers. Detectable Napsin TCRs were present in pre-treatment PBMC from both *A*02* carriers (14 of 27; 52%) and non-*A*02* carriers (9 of 28; 32%). The presence of Napsin TCRs at pre-treatment timepoints was associated with significantly improved PFS and OS for *A*02* carriers, (median PFS 21.5 months for Napsin TCR detectable vs. 7.2 months for Napsin TCR undetectable; p=0.031 and HR 0.37, median OS 60.2 months for Napsin TCR detectable vs. 16.5 months for Napsin TCR undetectable; p=0.0054 and HR 0.24 **Figure 3**). In contrast, there was no significant difference in PFS or OS based on the presence (n=9) or absence (n=19) of Napsin TCRs among non-*A*02* carriers (median PFS 6.3 months for Napsin TCR detectable vs. 5.2 months for Napsin TCR undetectable, p=0.94; median OS 25.8 months for Napsin TCR detectable vs. 19.1 months for Napsin TCR undetectable, p=0.21).

**Figure 3:**
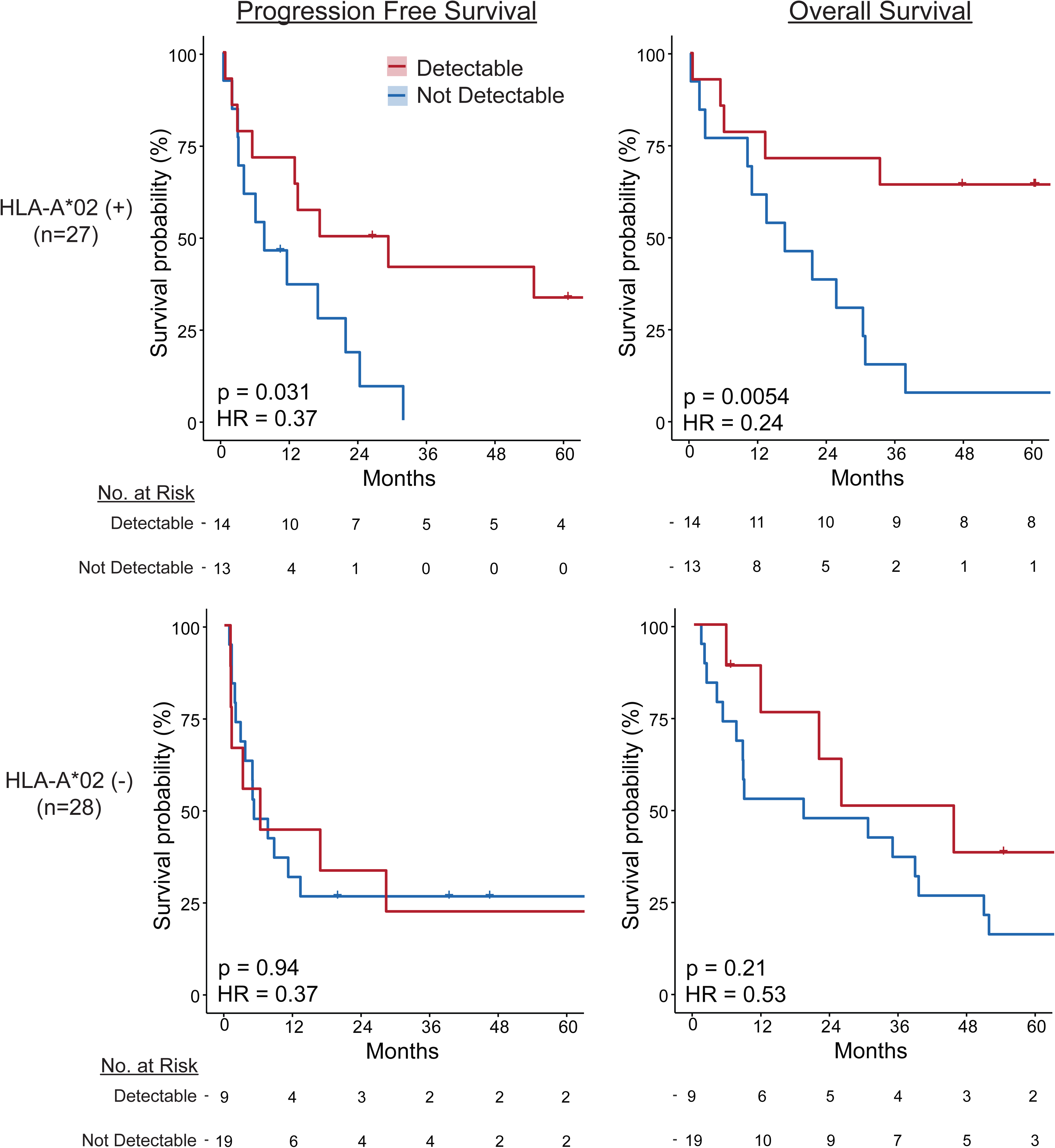
Improved PFS and OS among *HLA-A02*+ patients with detectable versus undetectable Napsin TCRs. Patients were separated into *HLA-A*02* carriers (n=27) and non-*HLA-A*02* carriers (n=28). Within each cohort, PFS and OS were plotted by Kaplan Meier method for patients with detectable Napsin TCRs (red) versus those without Napsin TCRs (blue). Comparisons were made using log-rank test. Among *HLA-A*02*(+) patients, those with Napsin TCRs detectable in pre-treatment PBMC (n=14) had significantly improved PFS and OS compared to those without (n=13; p=0.031 for PFS, p=0.0054 for OS. In contrast, among *HLA-A*02*(-) patients there was no significant difference between PFS or OS based on the presence (n=9) or absence (n=19) of Napsin TCRs (p=0.94 for PFS; p=0.21 for OS). A similar trend was observed among post-treatment samples, which was not statistically significant.

### Presence of Napsin TCRs as an independent variable associated with improved OS in univariate and multivariate analysis

To further compare Napsin TCRs to variables with established and/or potential value in predicting ICI response, we assessed the associations between select clinical characteristics and TCR variables with either OS or PFS using Cox proportional hazards regression (**Figure 4; Supplemental Table 2**). Clinical characteristics included previously identified metrics of improved response to ICI, including smoking status (current/former vs never), PD-L1 expression (≥ 50% vs <50%), histology (squamous vs adenocarcinoma); as well as age (≥ 70 vs <70), gender, ECOG performance status (0 vs ≥ 1), and history of IRAE (yes/no). Continuous variables included white blood count (WBC), absolute neutrophil count (ANC), absolute lymphocyte count (ALC), and albumin; line of therapy was retained as an ordinal variable (1st-5th). TCR metrics included Napsin A TCRs (binary as detectable vs not detectable), TCR richness, productive clonality, and fraction of T cells of all nucleated cells as continuous variables.

**Figure 4:**
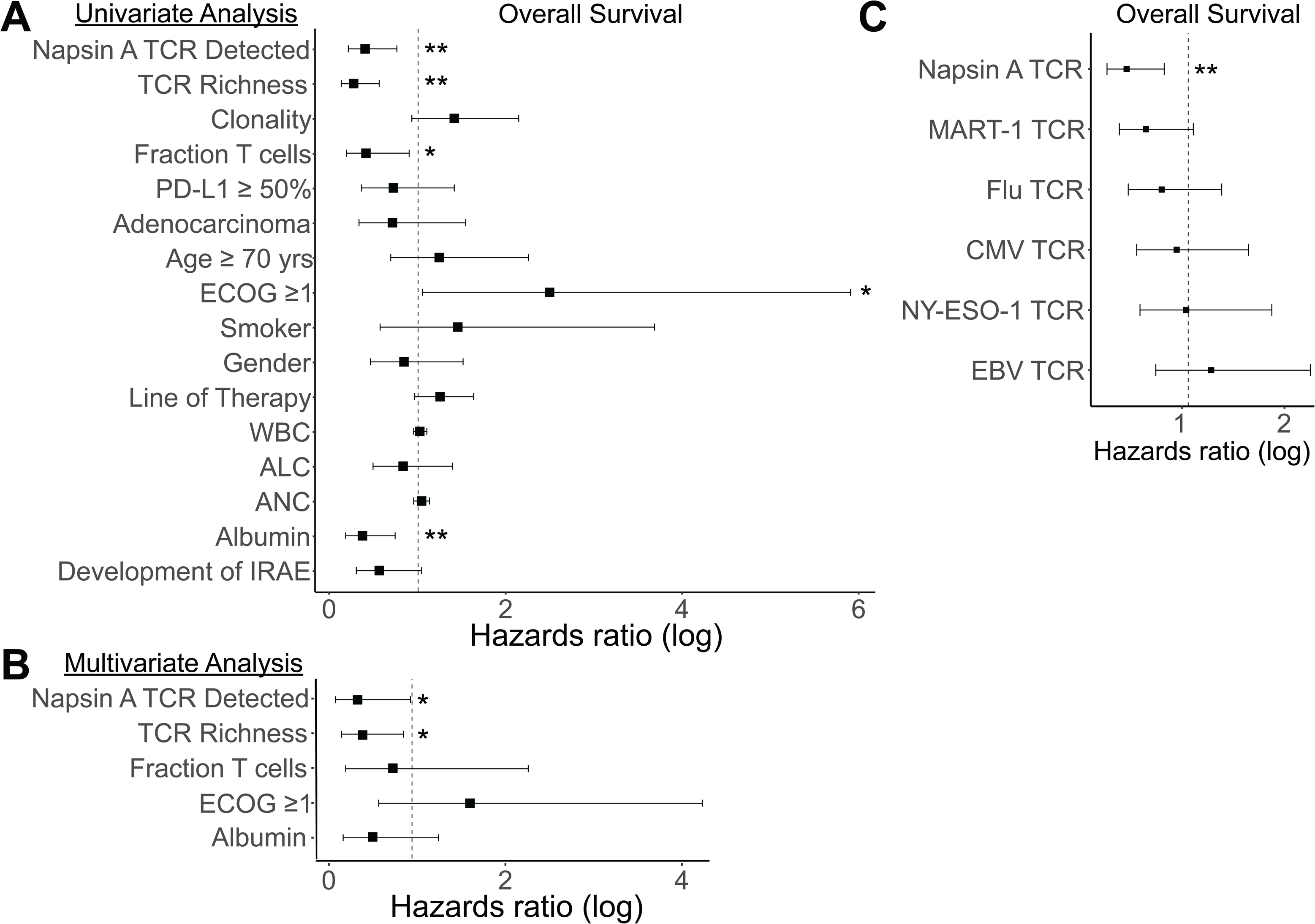
Presence of Napsin TCRs is associated with improved OS in univariate and multivariate analysis. Forest plot showing Hazard Ratio (HR) on log scale for each characteristic and association with OS by A) univariate analysis of TCR repertoire and clinical characteristics; B) multivariate analysis modeling association of characteristics significant from univariate analysis with OS. Cox proportional hazards regression analysis, values < 1 represent improved survival with presence of the indicated characteristic and C) univariate analysis with frequency of Napsin A- and other antigen-specific TCR above median (for Napsin A- and NY-ESO-1 median value = 0 thus above median is equivalent to Detected); * p < 0.05, ** p< 0.01.

Using univariate Cox regression analysis, detectable Napsin TCRs were associated with improved OS (p = 0.0057, HR 0.40, 95% CI 0.21-0.76) but not with PFS (p=0.17, HR 0.66, 95% CI 0.37-1.2; **Figure 4A** for OS**; Supplemental Figure 3** for PFS**; Supplemental Table 2** for complete statistical output). In addition, a higher fraction of T cells, albumen, and ECOG <1 were all associated with improved OS (Figure 4A).

Next, multivariate analysis was subsequently performed including all metrics found to be significantly associated with OS in univariate analysis (**Figure 4B**). Presence of Napsin TCRs was again significantly associated with improved OS (HR 0.45, 95% CI 0.23-0.91, p=0.025). A greater TCR richness was also associated with improved OS (HR 0.40, 95% CI 0.16-0.98, p=0.045), but fraction of T cells, ECOG, and higher albumin were not. The presence of Napsin TCRs remained a significant predictor of better OS, independent of TCR richness and other clinical characteristics and TCR variables.

Lastly, we assessed whether the frequency of other antigen-specific TCRs (beyond Napsin A) were associated with improved survival (**Figure 4C**). We determined the frequency of published TCRβ CDR3 sequences specific for epitopes from the lung-tropic viruses including influenza virus, EBV, and CMV as well as two tumor-associated antigens MART-1 (associated with melanoma) and NY-ESO-1 (associated with melanoma and a subset of ∼15% NSCLC ^22^) from public databases (**Supplemental Table 1**), and stratified patients using median TCR frequencies. Frequency of the antigen-specific TCR above the median was associated with improved OS for Napsin A only (p = 0.0057, HR 0.40, 95% CI 0.21-0.76) and not for the other five epitopes, consistent with an antigen-specific association.

### The frequency of Napsin TCRs is higher among patients with Durable Clinical Benefit (DCB)

We compared the frequencies of TCRs specific for Napsin A as well as other viral or tumor antigens among patients with or without DCB (**Table 2**). There was a significantly higher frequency of Napsin TCRs among patients with DCB compared to no durable clinical benefit (NDCB) (p = 0.025) in post-treatment PBMC (n=42; **Table 2**). A similar trend was observed from pre-treatment samples (n=62), which did not achieve statistical significance (p = 0.094). In comparison, the frequency of TCR specific for additional tumor antigens MART-1 and NY-ESO-1 or for viral antigens from CMV or EBV were not elevated among patients with DCB compared to patients without DCB. The frequency of TCRs specific for influenza virus was elevated among patients with DCB compared to patients with NDCB at pre-treatment timepoint only (p=0.024).

**Table 2:**
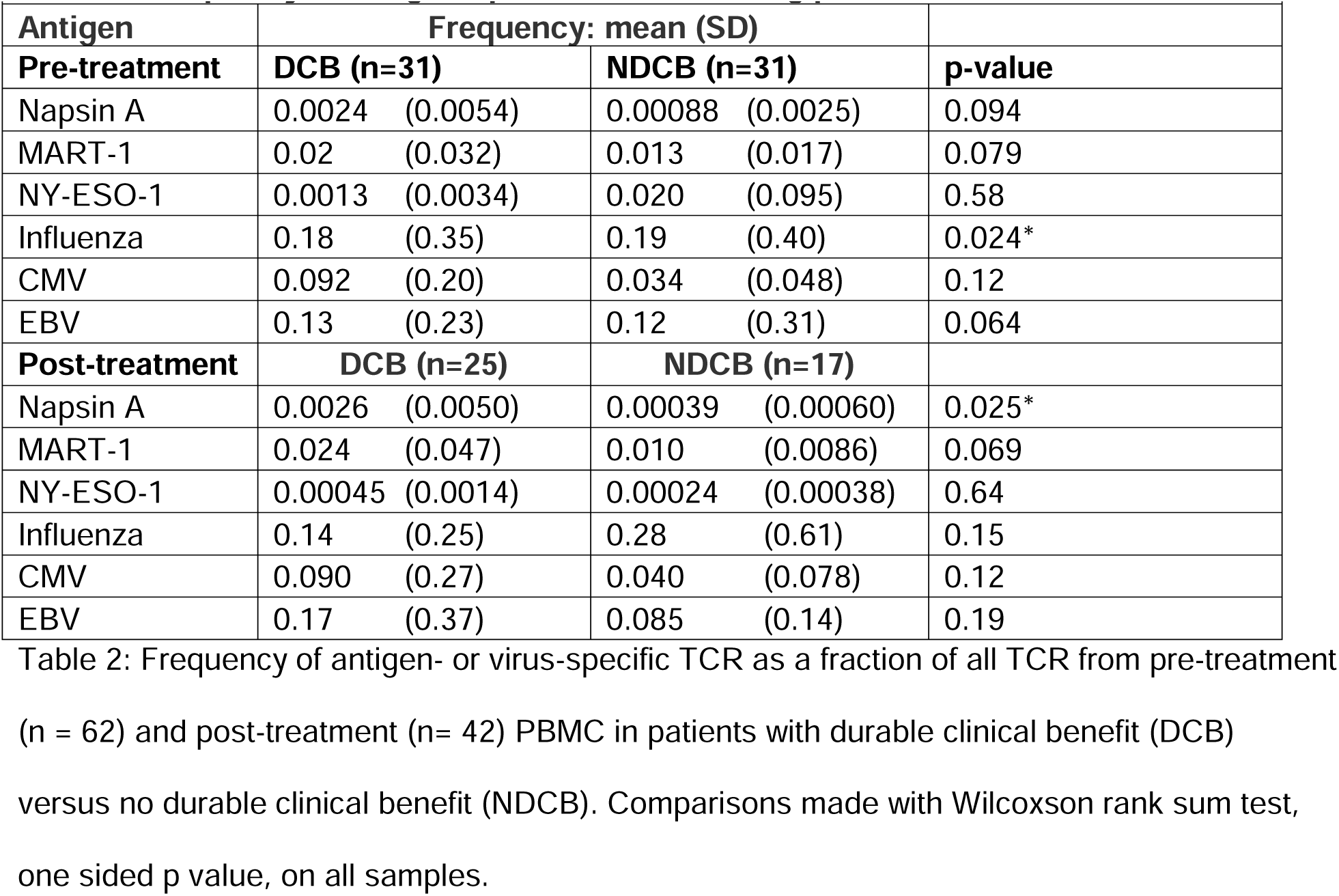
Frequency of antigen-specific T cells among patients with or without DCB.

### Some Napsin A TCRs have sequences identical to TCRs recognizing influenza antigens

We noted that many annotated CDR3β sequences specific for Napsin A were exact matches with CDR3β sequences that were also previously annotated to recognize the influenza A virus antigen M1 (Flu M1). Among the 20 Napsin TCRs, 11 CDR3β chain sequences were identical to CDR3β sequences identified as Flu M1-specific in publicly available databases (**Figure 5**). Of the patients with any of the 20 Napsin TCRs, one or more of these 11 ‘overlap’ TCRs were identified in 12 of 25 patients pre-treatment and 11 of 20 patients post-treatment. This finding implies that T cell recognition of Napsin A may be immunogenic due to its resemblance to Flu M1, despite being a self-antigen.

**Figure 5:**
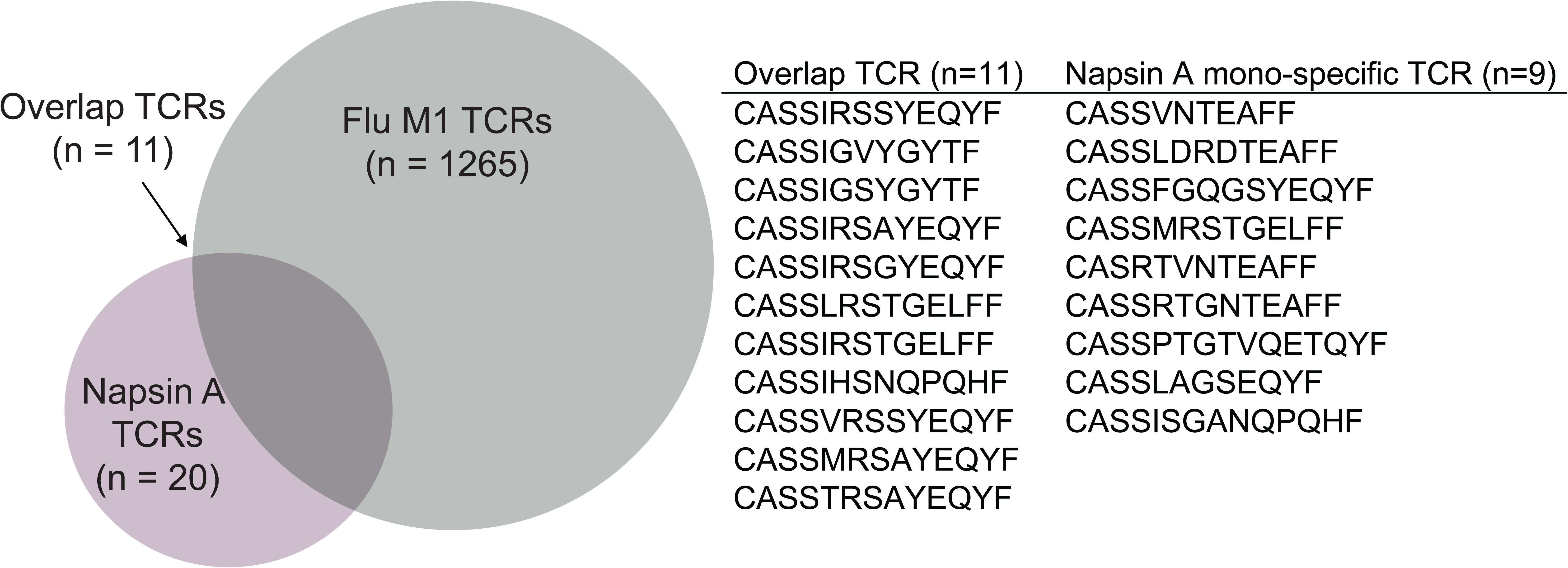
A significant fraction of Napsin TCRs are identical to known Flu M1-specific TCR and are enriched among patients responding to immunotherapy. Among TCR publicly annotated as specific for Flu M1 (n=1265; grey circle) and those specific for Napsin A (n=20; purple circle), there was a significant overlap of identical TCRβ CDR3 (n=11). The 20 Napsin specific TCR are shown, divided between those that overlap with Flu M1 (left) versus those that do not (n=9; right).

### No significant differences seen in tumor characteristics between patients with and without Napsin TCRs in the blood

To confirm that Napsin TCR detection in the blood was not related to underlying genomic features of the tumor, we assessed the tumor mutational profiles beyond the previously excluded *EGFR*, *ALK*, and/or *ROS1* alterations. We specifically assessed whether actionable genomic alterations in *KRAS*, *MET*, *RET*, *BRAF*, *ERBB2*/HER2, and/or *NTRK1/2/3* were different among patients with or without Napsin TCRs. Tumor mutational data was available for 54 of 62 patients (n= 22 with detectable Napsin TCRs, n=32 undetectable). Among patients with detectable Napsin TCRs at baseline, mutations included *KRAS* (p.G12C, n=2; non-G12C, n= 2), *MET* amplification (n=1), and *ERBB2* Exon 20 insertion (n=1). Among patients without detectable Napsin TCRs, mutations included *KRAS* (p.G12C, n=3; non-G12C n=7), *MET* exon 14 skipping mutation (n=1), and *ERBB2* Exon 20 insertion (n=1). There was no significant difference between the distribution of these mutations among patients with or without Napsin TCRs by Fisher’s exact test.

Next, we assessed whether there was an association between PD-L1 expression and Napsin TCRs at baseline. Baseline tumor PD-L1 expression was available for 51 of 62 patients. PD-L1 expression was ≥ 50% in seven of 21 with Napsin TCRs (33%) and 13 of 30 without (43%) (X^2^ = 0.51; p=0.47). We then evaluated whether presence of Napsin TCRs were restricted to any particular NSCLC histology. Napsin TCRs were detected at baseline in patients with all tumor histologies examined within our cohort at similar frequencies, including adenocarcinoma (n= 19 of 48 [40%]), SCC (n= 4 of 9 [44%]) and poorly differentiated NSCLC (n=2 of 5 [40%]). These findings support the hypothesis that Napsin TCRs reflect host immunity, and their presence in the blood is not associated with the genomic, molecular, or histologic characteristics of the tumor.

## DISCUSSION

The understanding of antigen-specific immune responses from checkpoint immunotherapy is limited and prior work to date has focused on neoantigen- or tumor-virus-specific responses ^1,2,5^. The extent to which overexpressed self-antigens in cancer play a role in ICI responses is less well established ^23^. Furthermore, TCR repertoire analysis have largely focused on descriptive metrics and insights into tumor antigen-specific immune responses have been limited due to the diversity of the TCR repertoire and the heterogeneity of the tumor antigens in lung cancer. Here we establish a dataset of TCR sequences from lung cancer patients treated with ICI across two institutions. We leverage publicly available sequences of TCRs targeting Napsin A, a widely overexpressed tumor antigen in lung adenocarcinoma, and quantify their frequencies in the peripheral blood of patients receiving ICI either as monotherapy or in combination therapy. We detected one or more of the 20 known Napsin TCRs in the peripheral blood in 25 out of 62 (40%) at the pre-treatment timepoint through TCR repertoire analysis. This is similar to the 39% of patients reported to have Napsin A-specific T cell responses in an independent cohort using orthogonal methods ^10^. We observed that patients with detectable Napsin TCRs (at both pre-treatment and post-treatment time points) had a significant improvement in OS, but not PFS, compared to patients without these TCRs. While cases of SCC were limited, the observed benefit of detectable Napsin TCRs on overall survival was only seen among patients with adenocarcinoma, not SCC; this is supportive of our hypothesis that detection of Napsin TCRs is associated with ICI response in NSCLC patients, particularly in lung adenocarcinoma histology where this antigen is highly expressed. When focusing survival analysis on *HLA-A*02* carriers, the likely HLA restriction for these TCRs isolated from four *HLA-A*02* carriers, there was significant improvement in both PFS and OS for patients with detectable Napsin TCRs. A previous report has demonstrated a trend toward Napsin A-specific T cell responses being associated with improved PFS and OS, as assayed by IFN-gamma release ^10^. This finding in an independent cohort using complementary methods does lend further support of our findings that detectable Napsin TCRs in the blood may be associated with durable immunity to checkpoint blockade.

Our results showed that detection of Napsin TCRs was associated with improved survival, independent of markers of general immune activation such as TCR richness, clonality, and/or fraction of T cells within a sample, and independent of clinical covariates known or suspected to impact outcomes such as performance status. In addition, we also determined that increased frequency of Napsin TCRs, but not TCR specific for lung-tropic viruses or other tumor associated antigens, was associated with improved overall survival. These data support the independent significance of Napsin A-specific T cells in our cohort.

We encountered the unexpected finding that many Napsin TCRs have also been previously annotated to be specific to the influenza M1 antigen. Despite being a self-antigen, Napsin A may be generating immunogenic peptides that can also be recognized by some Flu M1-targeting TCRs through molecular mimicry. In line with this, seasonal exposure to influenza antigens might play a role in the maintenance of Napsin TCRs

A limitation of our study is the heterogeneity of the patient population. However, the number of patients/samples in this study precluded further exploratory subset analyses. Another limitation of our study is inherent to TCR sequencing technology. Samples where Napsin TCRs are not detected may be because the reported sequences are truly not present, or because they are not detected by chance due to sampling. More broadly, we acknowledge that there are far more Napsin TCRs than the 20 assessed within this study and previously reported^10^ and that such TCRs may be specific to non *HLA-A*02* alleles. Therefore, the patients without detectable Napsin A-specific TCR within our study may indeed harbor alternative Napsin TCR not yet characterized. However, within these limitations, the 20 Napsin TCRs with this study appear to have immunogenic properties, based on their prevalence, frequencies, and sequence similarity to influenza-specific TCRs. Our study offers an analysis of TCR repertoire data that goes beyond descriptive metrics and interrogates antigen specificity focusing on Napsin A. Our study shows that monitoring the antigen-specific immune responses through TCR repertoire profiling can provide important insights into the antigenic determinants of checkpoint immunotherapy response.

In summary, T cells recognizing the self-antigen Napsin A may play a role in the durability of anti-tumor immune response to checkpoint immunotherapy. This suggests that it is not only neoantigens, but also potentially overexpressed self-antigens such as Napsin A, that can be immunogenic and among the determinants of response to checkpoint immunotherapy.

## Data Availability

All data produced in the present study are available upon reasonable request to the authors.

## DECLARATIONS

## Ethics approval and consent to participate

This study was approved by Fred Hutch Cancer Center IRB # 9358 and 6663/2242 and Stanford IRB #21319 (Molecular Analysis of Thoracic Malignancies)

## Consent for publication

All authors reviewed the manuscript and approved its publication.

## Availability of data and material

TCR sequencing data is available via the Adaptive Biotechnologies Published Projects Database and can be accessed at: https://clients.adaptivebiotech.com/pub/miller-2025-s.

## Authors’ contributions

NM, SC, DT, VN, and AMH were involved in study design. CB, JN, RS, SL, KN, RM, CR, HW, SP and DT treated the subject patients and were involved in sample/data collection. AK and TP isolated sample DNA for TCR sequencing. NM, DT, and FS performed chart review and data collection. NM and SC performed statistical analyses and generated figures. NM, SC and DT wrote the original manuscript with input from all authors who reviewed and approved the final manuscript.

## Acknowledgements

We acknowledge patients and families for participating in this research.

## List of Abbreviations

ALC: absolute lymphocyte count
ANC: absolute neutrophil count
CDR3: Complementarity-determining region 3
CMV: Cytomegalovirus
DCB: durable clinical benefit
EBV: Epstein–Barr virus
ECOG: Eastern Cooperative Oncology Group
FHCC: Fred Hutchinson Cancer Center
HLA: human leukocyte antigen
ICI: immune checkpoint inhibitor
IHC: immunohistochemistry
IRAE: immune related adverse event
MART-1: melanoma-associated antigen recognizing T cells 1
NDCB: non-durable clinical benefit
NSCLC: non-small cell lung cancer
NY-ESO-1: New York esophageal squamous cell carcinoma 1
OS: overall survival
PBMC: Peripheral blood mononuclear cells
PFS: progression-free survival
SCC: squamous cell carcinoma
TCR: T cell receptor
WBC: white blood count

## REFERENCES

1. Caushi JX, Zhang J, Ji Z, et al. Transcriptional programs of neoantigen-specific TIL in anti- PD-1-treated lung cancers [published correction appears in Nature. 2021 Oct;598(7881):E1. doi: 10.1038/s41586-021-03893-6]. Nature. 2021;596(7870):126–132. doi:10.1038/s41586-021-03752-4

2. Forde PM, Chaft JE, Smith KN, et al. Neoadjuvant PD-1 Blockade in Resectable Lung Cancer [published correction appears in N Engl J Med. 2018 Nov 29;379(22):2185. doi: 10.1056/NEJMx180040]. N Engl J Med. 2018;378(21):1976–1986. doi:10.1056/NEJMoa1716078

3. Komuro H, Shinohara S, Fukushima Y, et al. Single-cell sequencing on CD8^+^ TILs revealed the nature of exhausted T cells recognizing neoantigen and cancer/testis antigen in non- small cell lung cancer. J Immunother Cancer. 2023;11(8):e007180. doi:10.1136/jitc-2023-007180

4. Fehlings M, Jhunjhunwala S, Kowanetz M, et al. Late-differentiated effector neoantigen- specific CD8+ T cells are enriched in peripheral blood of non-small cell lung carcinoma patients responding to atezolizumab treatment. J Immunother Cancer. 2019;7(1):249. Published 2019 Sep 12. doi:10.1186/s40425-019-0695-9

5. Zhang J, Ji Z, Caushi JX, et al. Compartmental Analysis of T-cell Clonal Dynamics as a Function of Pathologic Response to Neoadjuvant PD-1 Blockade in Resectable Non-Small Cell Lung Cancer. Clin Cancer Res. 2020;26(6):1327–1337. doi:10.1158/1078-0432.CCR-19-2931

6. Reuben A, Zhang J, Chiou SH, et al. Comprehensive T cell repertoire characterization of non-small cell lung cancer. Nat Commun. 2020;11(1):603. doi:10.1038/s41467-019-14273-0

7. Weidemann S, Böhle JL, Contreras H, et al. Napsin A Expression in Human Tumors and Normal Tissues. Pathol Oncol Res. 2021;27:613099. Published 2021 Apr 20. doi:10.3389/pore.2021.613099

8. Tatnell PJ, Powell DJ, Hill J, Smith TS, Tew DG, Kay J. Napsins: new human aspartic proteinases. Distinction between two closely related genes. FEBS Lett. 1998;441(1):43–48. doi:10.1016/s0014-5793(98)01522-1

9. Turner BM, Cagle PT, Sainz IM, Fukuoka J, Shen SS, Jagirdar J. Napsin A, a new marker for lung adenocarcinoma, is complementary and more sensitive and specific than thyroid transcription factor 1 in the differential diagnosis of primary pulmonary carcinoma: evaluation of 1674 cases by tissue microarray. Arch Pathol Lab Med. 2012;136(2):163–171. doi:10.5858/arpa.2011-0320-OA

10. Berner F, Bomze D, Lichtensteiger C, et al. Autoreactive napsin A-specific T cells are enriched in lung tumors and inflammatory lung lesions during immune checkpoint blockade. Sci Immunol. 2022;7(75):eabn9644. doi:10.1126/sciimmunol.abn9644

11. Robins HS, Campregher PV, Srivastava SK, et al. Comprehensive assessment of T-cell receptor beta-chain diversity in alphabeta T cells. Blood. 2009;114(19):4099–4107. doi:10.1182/blood-2009-04-217604

12. Kirsch I, Vignali M, Robins H. T-cell receptor profiling in cancer. Mol Oncol. 2015;9(10):2063–2070. doi:10.1016/j.molonc.2015.09.003

13. Shannon, C.E. (1948) A Mathematical Theory of Communication. Bell System Technical Journal, 27, 379–423. 10.1002/j.1538-7305.1948.tb01338.x

14. Morisita M. Iσ-index, a measure of dispersion of individuals. Res Popul Ecol. 1962;4:1–7. 10.1007/BF02533903.

15. Shugay M, Bagaev DV, Zvyagin IV, et al. VDJdb: a curated database of T-cell receptor sequences with known antigen specificity. Nucleic Acids Res. 2018;46(D1):D419–D427. doi:10.1093/nar/gkx760

16. Yang X, Chen G, Weng NP, Mariuzza RA. Structural basis for clonal diversity of the human T-cell response to a dominant influenza virus epitope. J Biol Chem. 2017;292(45):18618–18627. doi:10.1074/jbc.M117.810382

17. Glanville J, Huang H, Nau A, et al. Identifying specificity groups in the T cell receptor repertoire. Nature. 2017;547(7661):94–98. doi:10.1038/nature22976

18. Pritchard CC, Salipante SJ, Koehler K, et al. Validation and implementation of targeted capture and sequencing for the detection of actionable mutation, copy number variation, and gene rearrangement in clinical cancer specimens. J Mol Diagn. 2014;16(1):56–67. doi:10.1016/j.jmoldx.2013.08.004

19. Altan M, Li R, Li Z, et al. High peripheral T cell diversity is associated with lower risk of toxicity and superior response to dual immune checkpoint inhibitor therapy in patients with metastatic NSCLC. J Immunother Cancer. 2024;12(12):e008950. doi:10.1136/jitc-2024-008950

20. Dong N, Moreno-Manuel A, Calabuig-Fariñas S, et al. Characterization of Circulating T Cell Receptor Repertoire Provides Information about Clinical Outcome after PD-1 Blockade in Advanced Non-Small Cell Lung Cancer Patients. Cancers. 2021; 13(12):2950. 10.3390/cancers13122950

21. Abed A, Beasley AB, Reid AL, et al. Circulating pre-treatment T-cell receptor repertoire as a predictive biomarker in advanced or metastatic non-small-cell lung cancer patients treated with pembrolizumab alone or in combination with chemotherapy. ESMO Open. 2023;8(6):102066. doi:10.1016/j.esmoop.2023.102066

22. Barnes B, Shan M, Blouch K, et al. Analysis of NY-ESO-1 expression in specimens from a Phase I/II NY-ESO-1 T-cell therapy clinical trial in non-small cell lung cancer and from exploratory studies in multiple tumor types. Journal for ImmunoTherapy of Cancer 2021;9: Abstract 454. doi:10.1136/jitc-2021-SITC2021.454

23. Oliveira G, Stromhaug K, Klaeger S, et al. Phenotype, specificity and avidity of antitumour CD8^+^ T cells in melanoma. Nature. 2021;596(7870):119–125. doi:10.1038/s41586-021-03704-y

