## Supplemental Table 1 for "Napsin A-specific T cell clonotypes are associated with improved clinical outcomes in patients receiving checkpoint immunotherapy for metastatic non-small cell lung cancer"

Supplemental Table 1: HLA-A typing of patients with sufficient genomic DNA (n=55)

| Patient | HLA-A allele 1 | HLA-A allele 2 |
|---|---|---|
| FHCC-A002 | A*02:01 | A*03:01 |
| FHCC-A005 | A*01:01 | A*03:01 |
| FHCC-A009 | A*02:01 | A*02:01 |
| FHCC-A014 | A*02:06 | A*02:01 |
| FHCC-A015 | A*01:01 | A*25:01 |
| FHCC-A016 | A*02:01 | A*29:02 |
| FHCC-A017 | A*02:01 | A*30:02 |
| FHCC-A018 | A*31:01 | A*31:01 |
| FHCC-A020 | A*02:01 | A*11:01 |
| FHCC-A022 | A*01:01 | A*02:01 |
| FHCC-A024 | A*01:01 | A*24:02 |
| FHCC-A025 | A*01:01 | A*23:01 |
| FHCC-A026 | A*02:01 | A*32:01 |
| FHCC-A028 | A*11:01 | A*24:02 |
| FHCC-A030 | A*03:01 | A*68:01 |
| FHCC-A032 | A*03:01 | A*03:01 |
| FHCC-A033 | A*29:02 | A*31:01 |
| FHCC-A035 | A*02:01 | A*68:01 |
| FHCC-A039 | A*02:01 | A*02:01 |
| FHCC-A040 | A*01:01 | A*32:01 |
| FHCC-A041 | A*03:01 | A*24:02 |
| FHCC-A042 | A*02:01 | A*23:01 |
| FHCC-A043 | A*11:01 | A*24:02 |
| FHCC-A047 | A*30:02 | A*34:02 |
| FHCC-A048 | A*01:01 | A*31:01 |
| FHCC-A050 | A*02:01 | A*29:02 |
| FHCC-A052 | A*02:01 | A*29:02 |
| FHCC-A054 | A*03:01 | A*31:01 |
| FHCC-A055 | A*02:01 | A*30:01 |
| FHCC-A056 | A*30:02 | A*32:01 |
| FHCC-A058 | A*02:01 | A*24:03 |
| FHCC-A059 | A*01:01 | A*02:01 |
| FHCC-A060 | A*02:01 | A*24:02 |
| FHCC-A061 | A*01:01 | A*68:01 |
| FHCC-A065 | A*11:01 | A*30:01 |
| FHCC-A066 | A*02:01 | A*24:02 |
| FHCC-A067 | A*02:01 | A*32:01 |
| FHCC-A068 | A*01:01 | A*26:01 |
| FHCC-A072 | A*02:01 | A*32:01 |
| FHCC-A073 | A*01:01 | A*29:02 |
| FHCC-A076 | A*02:01 | A*11:01 |

| | | |
|---|---|---|
| FHCC-A077 | A*02:01 | A*03:01 |
| LNGTCR015 | A*01:01 | A*02:01 |
| LNGTCR016 | A*11:01 | A*11:02 |
| LNGTCR020 | A*02:01 | A*32:01 |
| LNGTCR026 | A*01:01 | A*33:03 |
| LNGTCR028 | A*01:01 | A*26:01 |
| LNGTCR029 | A*11:01 | A*31:01 |
| LNGTCR035 | A*02:01 | A*03:01 |
| LNGTCR037 | A*01:01 | A*68:01 |
| LNGTCR042 | A*02:01 | A*02:24 |
| LNGTCR043 | A*24:02 | A*33:01 |
| LNGTCR047 | A*02:03 | N/A |
| LNGTCR056 | A*68:01 | A*68:02 |
| LNGTCR069 | A*26:01 | A*03:01 |
