## Supplemental Table 2 for "Napsin A-specific T cell clonotypes are associated with improved clinical outcomes in patients receiving checkpoint immunotherapy for metastatic non-small cell lung cancer"

Supplemental Table 2: Presence of Napsin A-specific TCR is associated with improved OS in univariate and multivariate analysis

| Univariate Analysis | Overall Survival | | | Progression Free Survival | | |
|---|---|---|---|---|---|---|
| | HR | 95% CI | p value | HR | 95% CI | p value |
| Napsin A TCR Detected | 0.40 | 0.21 - 0.76 | 0.0057** | 0.66 | 0.37 - 1.2 | 0.17 |
| TCR Richness | 0.27 | 0.13 - 0.56 | 0.00039** | 0.41 | 0.20-0.81 | 0.011* |
| Clonality | 1.4 | 0.93 - 2.1 | 0.10 | 1.1 | 0.72 – 1.7 | 0.62 |
| Fraction T cells | 0.41 | 0.19 - 0.90 | 0.025* | 0.51 | 0.23 – 1.1 | 0.093 |
| PD-L1 ≥ 50% | 0.72 | 0.36 - 1.4 | 0.34 | 0.69 | 0.36 – 1.3 | 0.27 |
| Adenocarcinoma | 0.71 | 0.33 - 1.5 | 0.39 | 0.90 | 0.42 – 2.0 | 0.80 |
| Age ≥ 70 | 1.2 | 0.69 - 2.3 | 0.48 | 1.1 | 0.64 – 2.0 | 0.67 |
| ECOG ≥ 1 | 2.5 | 1.0 - 5.9 | 0.039* | 2.9 | 1.2 – 6.8 | 0.016* |
| Smoker | 1.5 | 0.57 - 3.7 | 0.43 | 1.2 | 0.51 - 2.8 | 0.68 |
| Male Gender | 0.84 | 0.46 - 1.5 | 0.55 | 0.77 | 0.43 – 1.4 | 0.37 |
| Line of Therapy | 1.3 | 0.96 - 1.6 | 0.097 | 1.2 | 0.92 – 1.6 | 0.18 |
| WBC | 1.0 | 0.95 - 1.1 | 0.61 | 1.0 | 0.95 – 1.1 | 0.63 |
| ALC | 0.83 | 0.49 - 1.4 | 0.48 | 0.77 | 0.46 – 1.3 | 0.30 |
| ANC | 1 | 0.95 - 1.1 | 0.39 | 1.0 | 0.96 – 1.1 | 0.34 |
| Albumin | 0.37 | 0.18 - 0.74 | 0.0051** | 0.38 | 0.19 – 0.74 | 0.0042** |
| Development of IRAE | 0.56 | 0.30 - 1.0 | 0.064 | 0.60 | 0.33 1.108 | 0.086 |

| Multivariate Analysis | Overall Survival | | |
|---|---|---|---|
| | HR | 95% CI | p value |
| TCR Richness | 0.40 | 0.16 - 0.98 | 0.045* |
| Napsin A TCR Detected | 0.45 | 0.23 - 0.91 | 0.025* |
| Fraction T cells | 0.79 | 0.27 - 2.3 | 0.66 |
| ECOG ≥ 1 | 1.6 | 0.63 - 4.2 | 0.31 |
| Albumin | 0.56 | 0.24 - 1.3 | 0.17 |

| Univariate Analysis | Overall Survival | | |
|---|---|---|---|
| | HR | 95% CI | p value |
| Napsin A TCR | 0.40 | 0.21 - 0.76 | 0.0057** |
| MART-1 TCR | 0.59 | 0.33 – 1.1 | 0.072 |
| Influenza TCR | 0.74 | 0.41 – 1.3 | 0.31 |
| CMV TCR | 0.89 | 0.50 - 1.6 | 0.69 |
| NY-ESO-1 TCR | 0.98 | 0.53 - 1.8 | 0.95 |
| EBV TCR | 1.2 | 0.68 – 2.2 | 0.50 |
