## Supplemental Figure 1 for "Napsin A-specific T cell clonotypes are associated with improved clinical outcomes in patients receiving checkpoint immunotherapy for metastatic non-small cell lung cancer"

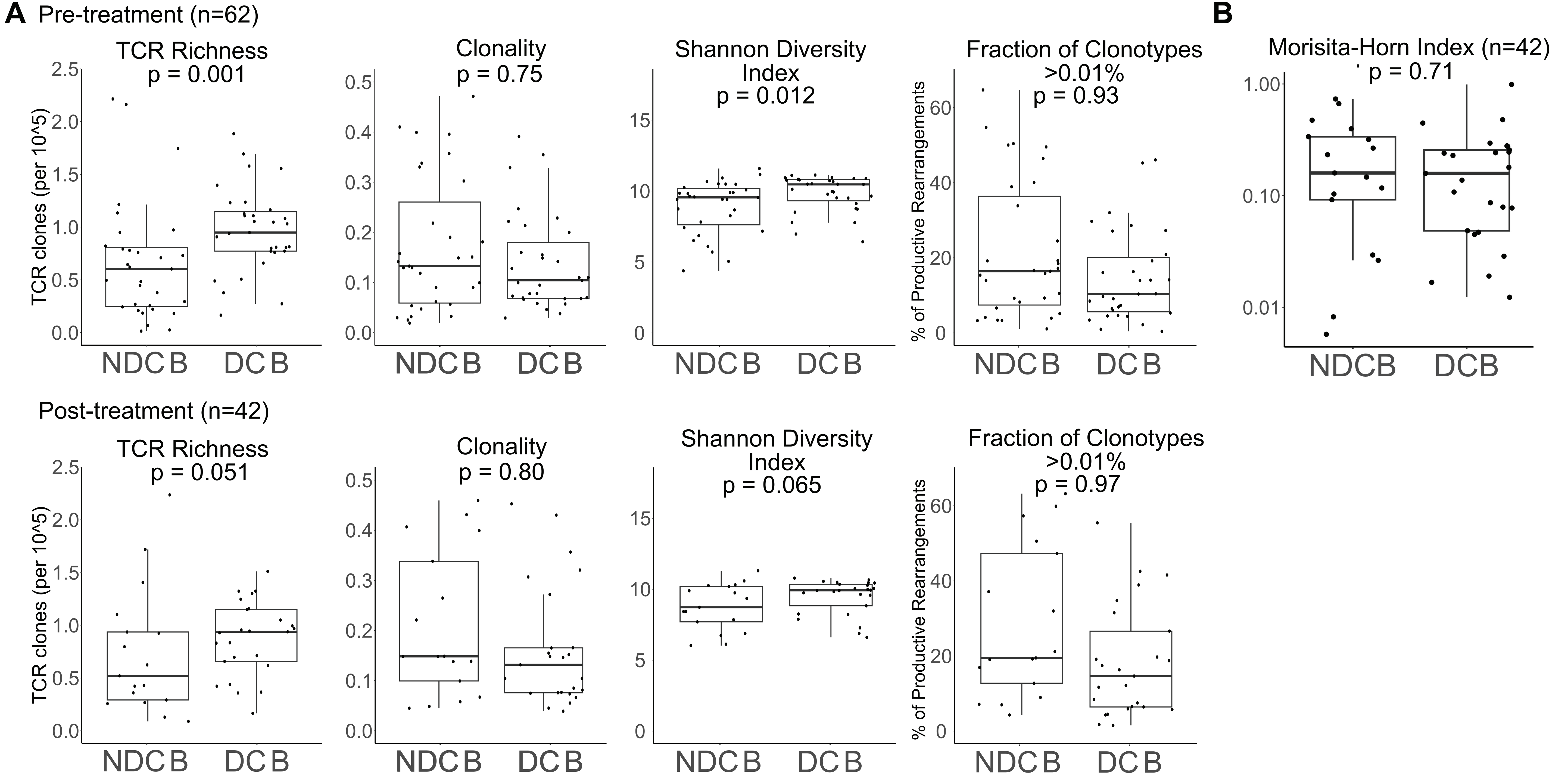


**Supplemental Figure 1:** A) Metrics of TCR repertoire from PBMC obtained at pre-treatment (n = 62) and post-treatment (n= 42) timepoints in patients with durable clinical benefit (DCB) versus no durable clinical benefit (NDCB). B) Level of similarity between pre- and post- treatment TCR repertoires within each patient per Morisita-Horn similarity index (scale 0-1; with values toward 1 indicating high correlation with many TCR at similar frequencies and 0 indicating no shared TCRs), grouped by clinical benefit. Comparisons made with Wilcoxson rank sum test, one sided p-value, on all samples. Box plots illustrate median of non-zero values with 25th and 75th percentile quantiles.
