## Supplemental Figure 2 for "Napsin A-specific T cell clonotypes are associated with improved clinical outcomes in patients receiving checkpoint immunotherapy for metastatic non-small cell lung cancer"

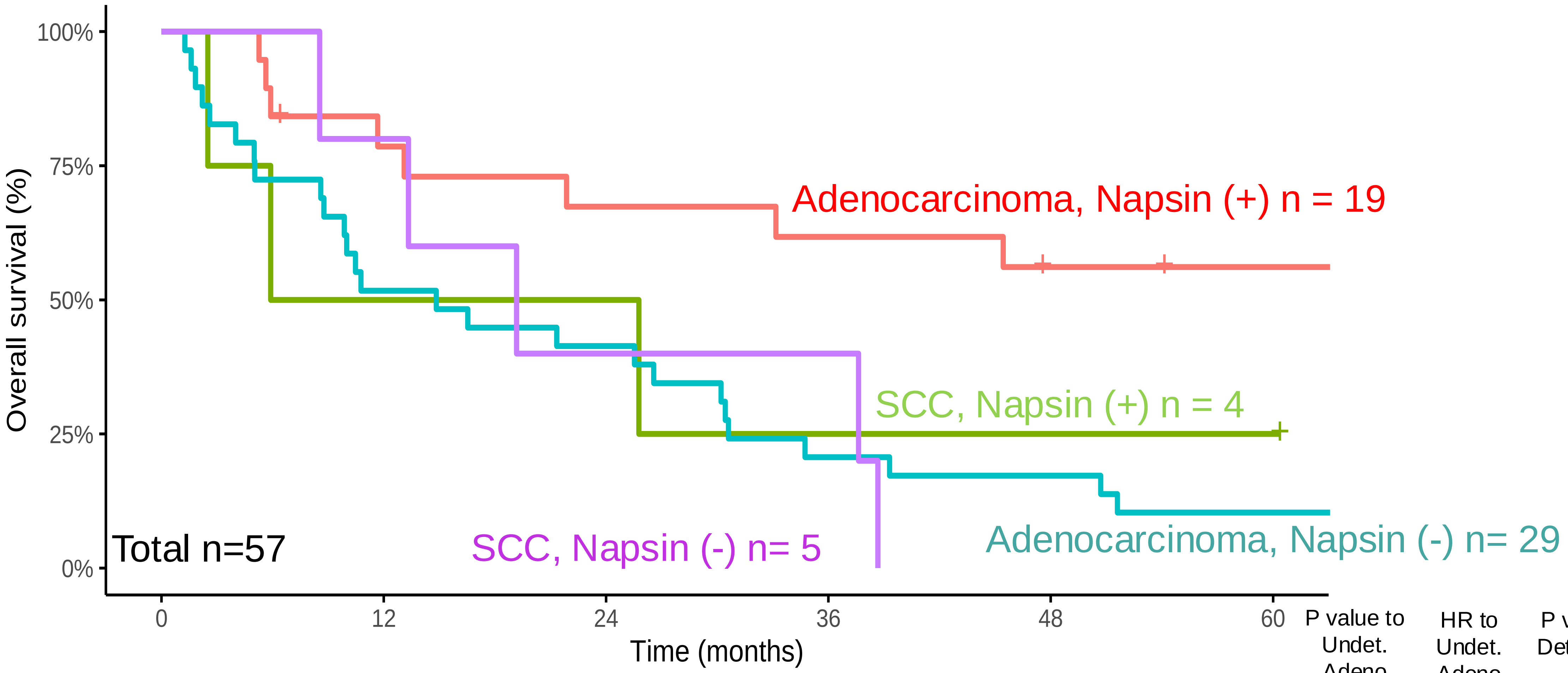


| | 0 | 12 | 24 | 36 | 48 | 60 | P value to Undet. Adeno | HR to Undet. Adeno | P value to Detectable SCC |
|---|---|---|---|---|---|---|---|---|---|
| Detectable Adeno | 19 | 14 | 12 | 11 | 9 | 8 | 0.0019 | 0.30 | - |
| Detectable SCC | 4 | 2 | 2 | 1 | 1 | 1 | - | - | - |
| Undetectable Adeno | 29 | 15 | 12 | 6 | 5 | 3 | - | - | - |
| Undetectable SCC | 5 | 4 | 2 | 2 | 0 | 0 | - | - | 0.90 |

**Supplemental Figure 2:** Overall survival among patients with adenocarcinoma or squamous cell carcinoma (SCC) histology (n=57), excluding those with poorly differentiated NSCLC (n= 5). Kaplan-Meier method used to plot survival curves categorized by histology and presence or absence of Napsin A-specific TCR in pre-treatment PBMC. Number at risk at each timepoint shown below plot. Log-rank test used for comparisons. Adenocarcinoma patients with detectable Napsin-A specific TCR had improved OS compared to adenocarcinoma patients without Napsin A-specific TCR (n = 0.002; HR 0.30); in contrast no significant OS difference was seen among SCC patients with or without detectable Napsin A-specific TCR (p=0.9).
