## Supplemental Figure 3 for "Napsin A-specific T cell clonotypes are associated with improved clinical outcomes in patients receiving checkpoint immunotherapy for metastatic non-small cell lung cancer"

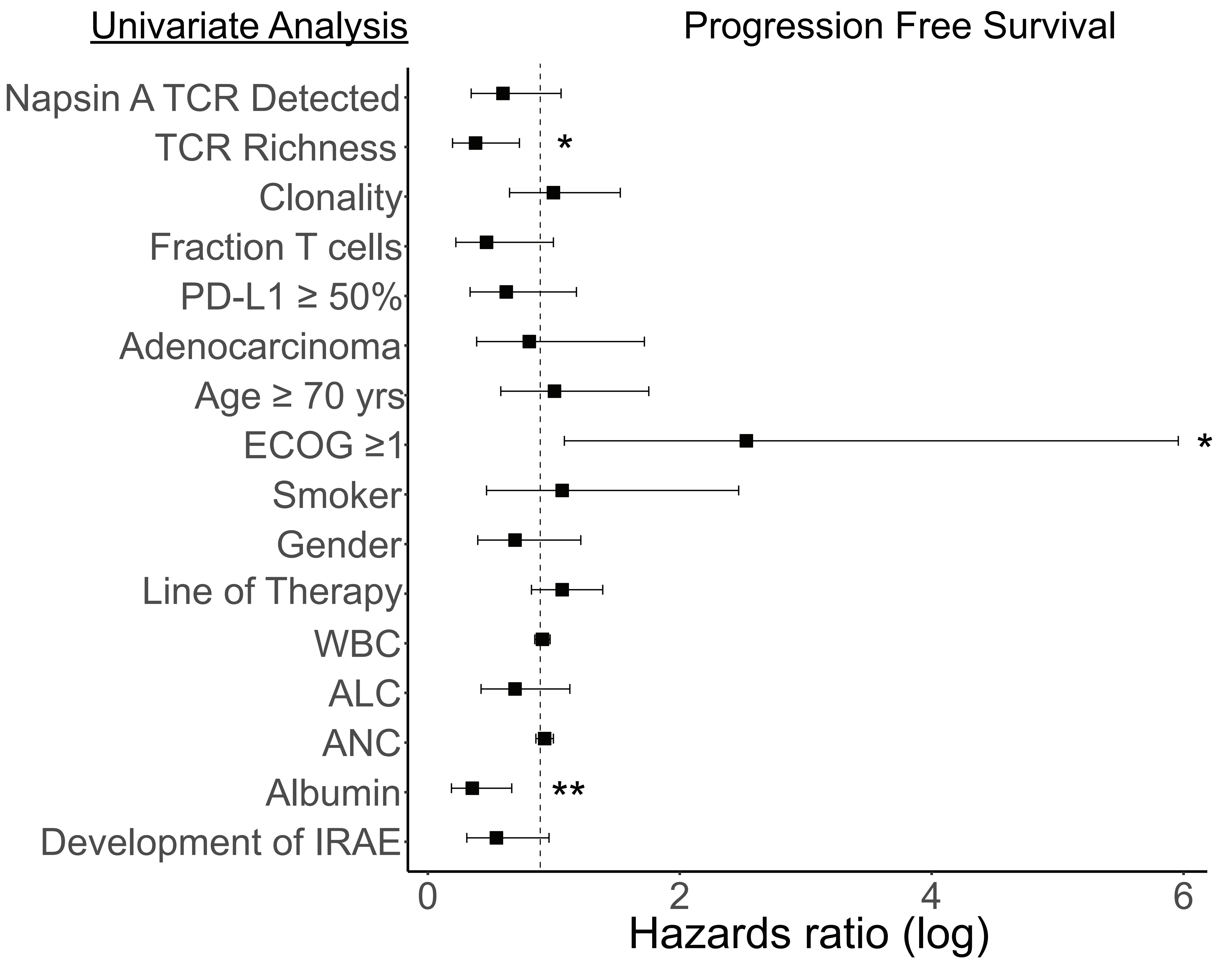


**Supplemental Figure 3:** Presence of Napsin A-specific TCR is not associated with improved PFS in univariate analysis. Forest plot showing Hazard Ratio (HR) on log scale for each characteristic and association with PFS. Cox proportional hazards regression analysis, values $< 1$ represent improved PFS with presence of the indicated characteristic.* $p < 0.05$, ** $p < 0.01$
